# Combination Venetoclax and Selinexor Effective in Relapsed Refractory Multiple Myeloma with Translocation t(11;14)

**DOI:** 10.1101/2022.08.01.22278282

**Authors:** Nina Nguyen, Sana Chaudhry, Tulasigeri M. Totiger, Robert Diaz, Evan Roberts, Skye Montoya, Gabriel Pardo, Alejandro Pardo, Jumana Afaghani, Maurizio Affer, Jacob Jahn, Terrence Bradley, Francesco Maura, Dickran Kazandjian, Daniel Bilbao, Jennifer Chapman, Ola Landgren, James Hoffman, Justin Taylor

## Abstract

Patients with multiple myeloma bearing translocation t(11;14) have recently been shown to benefit from the apoptosis-inducing drug venetoclax; however, the drug lacks FDA approval in multiple myeloma thus far due to a potential safety signal in the overall patient population. Selinexor is an inhibitor of nuclear export that is FDA-approved for patients with multiple myeloma refractory multiple lines of therapy. Here, we report that in four patients with multiple myeloma with t(11;14), the concomitant administration of venetoclax and selinexor was safe and associated with disease response. Moreover, the combination was synergistic in t(11;14) multiple myeloma cell lines and caused decreased levels of Cyclin D1 (which is overexpressed due to the *CCND1-IGH* fusion) when given in combination as compared to single agents. These data suggest that the combination of venetoclax and selinexor is effective and t(11;14) may serve as a therapeutic marker for response and target for future clinical trials.

## Introduction

In the t(11;14)(q13;q32) translocation, *CCND1* at chromosome 11q13 is juxtaposed to the *IGH* gene at chromosome 14q32, resulting in overexpression of the protein Cyclin D1.^1–4^ Screening of multiple myeloma (MM) cell lines for sensitivity to venetoclax showed that high sensitivity was restricted to cell lines with *CCND1* translocations.^5,6^ Venetoclax induces apoptosis by acting as a BH3-mimetic to inhibit the anti-apoptotic factor BCL2^7^ and is approved for use in certain B-cell malignancies and acute myeloid leukemia. Venetoclax has been studied in combination with bortezomib and dexamethasone (versus placebo with bortezomib and dexamethasone) for relapsed/refractory myeloma; however, the benefit in progression-free survival (PFS) in the venetoclax arm was offset by increased mortality, thus hindering its path to approval. A subset analysis showed that patients who possessed t(11;14) and/or high BCL2 expression had clinical responses and longer PFS without increased mortality^8^, leading to cautious off-label use of the medication in select patients. This benefit was confirmed in the final trial analysis after longer follow-up.^9^ Selinexor is an inhibitor of nuclear export (SINE) that blocks the cargo-binding pocket of XPO1. XPO1 recognizes cargo proteins bearing a nuclear export signal and shuttles them out of the nucleus. These cargoes include tumor suppressor proteins such as p53, as well as proteins binding to select mRNA including BCL2.^10^ Cyclin D1 is a known cargo protein of XPO1.^11,12^ Cyclin D1 is essential in regulation of the cell cycle, and its overexpression can result in uncontrolled cell growth, contributing to cancer development and progression.^13–15^ Selinexor is approved for patients with myeloma who have received at least four prior lines of therapy and are refractory to two proteasome inhibitors, two immunomodulating therapies, and a CD38-targeted monoclonal antibody.^16–18^

We report here, four patients with relapsed/refractory t(11;14) MM who had progression of disease after multiple lines of treatment and were considered for venetoclax treatment based on previous data showing efficacy of venetoclax in t(11;14) MM.^8,19^ All patients responded initially to venetoclax but ultimately developed resistance and progressive disease. The addition of selinexor recaptured responses and lead to clinical benefit, suggesting a synergistic effect of the combination. The combination of venetoclax and selinexor was further studied in MM cell lines with and without t(11;14) translocations and showed enhanced synergy in those cell lines bearing the *CCND1*-*IGH* translocation.

## Results

### Patients with relapsed and refractory multiple myeloma with t(11;14) tolerate and respond to selinexor and venetoclax combination

#### Cases

A 55-60 year-old African-American man with free kappa light chain MM, R-ISS stage 1, and standard risk cytogenetics with t(11;14); (**Figure 1A**) was treated with selinexor and venetoclax off-label therapy. The patient had progression of disease (POD) through seven lines of therapy which are depicted in **Figure 1B**. The patient’s best response to therapy was a very good partial response (VGPR) with daratumumab, pomalidomide, and dexamethasone (dara-PD). He otherwise had minimal response (MR) to prior regimens. He completed 25 cycles of dara-PD with eventual POD. Due to lack of other therapeutic options, he received venetoclax 400mg daily and achieved a partial response (PR), then a VGPR with the addition of bortezomib 1.3 mg/m2 every two weeks and dexamethasone 12 mg weekly. The patient had 17 cycles of treatment before POD, at which point bortezomib was substituted for carfilzomib with continued POD. The patient was enrolled onto a clinical trial of a BCMAxCD3 bi-specific T-cell engager with POD. Given prior resistance to venetoclax-based therapy, he was given selinexor 60 mg weekly in combination with venetoclax 400 mg daily and dexamethasone 40 mg weekly with subsequent VGPR. His kappa light-chain levels correlated with response and are shown in **Figure 1C**. The patient had a 10-month duration of response. Upon progression, the patient was then treated with commercial chimeric antigen receptor T-cell (CAR-T) therapy.

**Figure 1.**
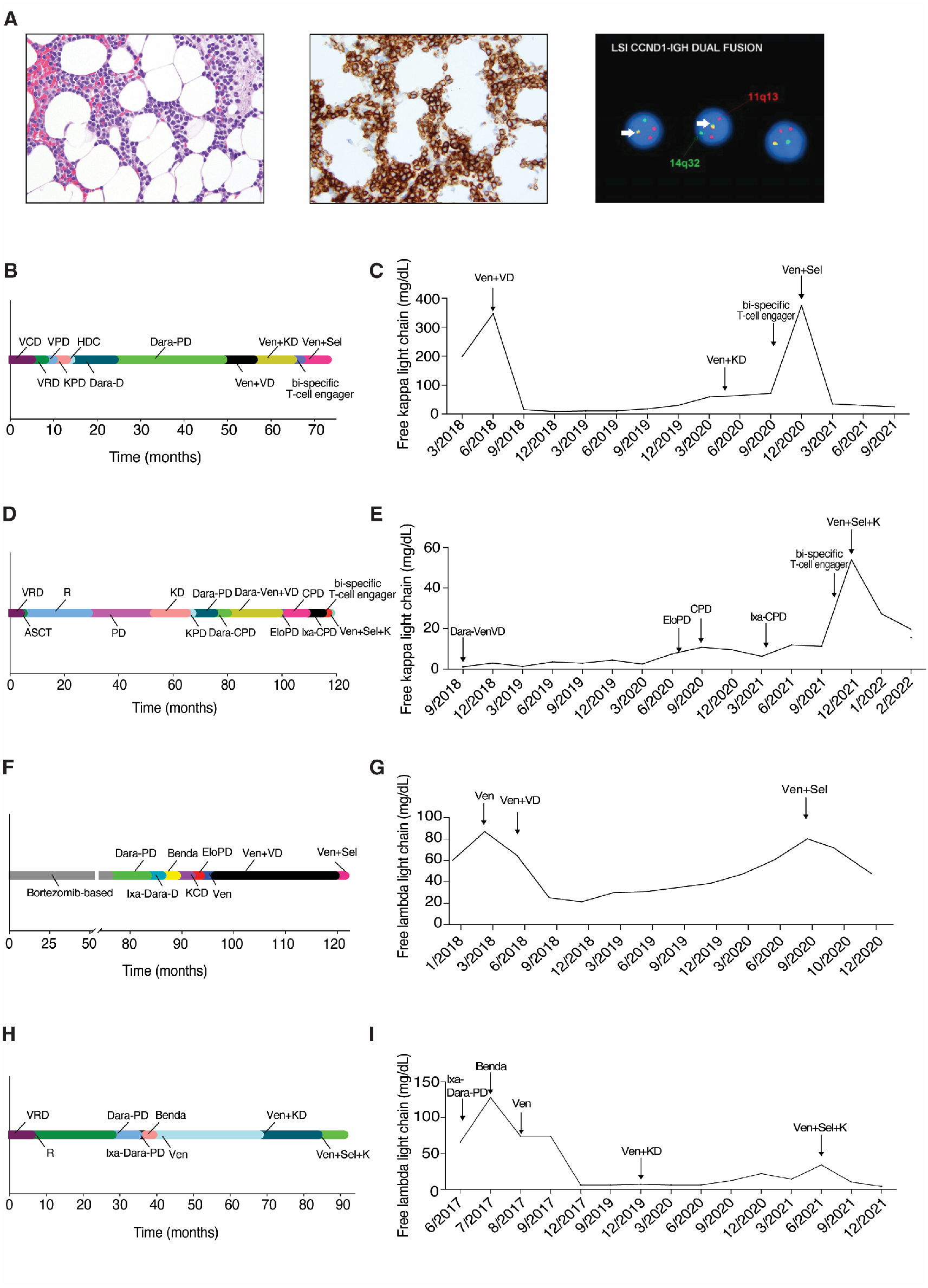
Clinical response to selinexor and venetoclax in heavily pre-treated t(11;14) multiple myeloma. Histopathologic findings in bone marrow biopsy at diagnosis showed extensive involvement by plasma cell myeloma. Neoplastic plasma cells were small and mature in appearance and present in a diffuse interstitial distribution comprising ∼50-60% of overall marrow cellularity. Hematoxylin and Eosin stain, 400x magnification (**Panel A left**). Immunohistochemistry (IHC) for CD138 highlights neoplastic plasma cells present in abnormal clusters, 400x magnification (**Panel A middle**). Fluorescence *in situ* hybridization (FISH) studies showed *CCND1*-*IGH* fusion in 95% of cells (yellow fusion signals indicated by arrow; **Panel A right**). Timeline of prior treatments and free light chain response for Patient 1 (**Panel B** and **Panel C**), Patient 2 (**Panel D** and **Panel E**), Patient 3 (**Panel F** and **Panel G**) and Patient 4 (**Panel H** and **Panel I**). Legend-V: bortezomib, C: cyclophosphamide, D: dexamethasone, R: lenalidomide, P: pomalidomide, K: carfilzomib, HDC: high dose cyclophosphamide, Dara: daratumumab, Ven: venetoclax, Sel: selinexor, Ixa: ixazomib, Elo: elotuzumab, ACST: autologous stem cell transplant, Benda: bendamustine

An additional three patients with t(11;14) MM were treated with the combination of selinexor and venetoclax. The summary of their treatment history and responses to therapy are outlined in **Figure 1**. Patient 2 had a PR to therapy with a duration of 4 months; patient 3 had a MR to therapy with a duration of 3 months; and patient 4 had a VGPR to therapy with a duration of 6 months. Patient 3 enrolled on clinical trial after progression on selinexor and venetoclax. Patients 2 and 4 are now being considered for CAR-T therapy due to progression of disease on the combination. We have treated a total of 4 patients with the combination of venetoclax and selinexor, and 3 patients had a PR or better.

Overall, the combination of selinexor and venetoclax was well tolerated. Selinexor was prescribed at 60 mg or 80 mg weekly, and venetoclax was prescribed at either 400 mg or 800 mg daily based upon tolerance and efficacy. One patient had dose interruption of selinexor due to fatigue and dyspnea, however she was able to resume therapy without further issues. No other dose interruptions or reductions were required. Leukopenia, neutropenia, and thrombocytopenia were observed, but there was no incidence of bleeding or febrile neutropenia. Hyponatremia was also observed, however did not require intervention and resolved.

### Multiple myeloma cell lines with t(11;14) show high levels of synergism between selinexor and venetoclax *in vitro*

To further test our hypothesis of the preferential effects of selinexor and venetoclax in patients bearing the t(11;14) translocation, we tested the synergy of the combination on human MM cell lines at various concentrations and then examined cell viability 72 hours after treatment. MM cell lines with and without the t(11;14) translocation are shown in **Supplemental Table 1**. The combination of selinexor and venetoclax showed synergy in all multiple myeloma cell lines tested (**Figure 2A-E** and **Fig. 1A-E** in the **Supplementary Appendix**). U266-B1, bearing the t(11;14) translocation, showed a higher level of synergy in comparison to the non-t(11;14) translocation cell line RPMI-8226 (**Figure 2C**). We tested two additional cell lines and saw similar results of higher level of synergy in the t(11;14) translocation cell line in comparison to the non-t(11;14) cell line (**Figure 1C** in the **Supplementary Appendix**).

**Figure 2.**
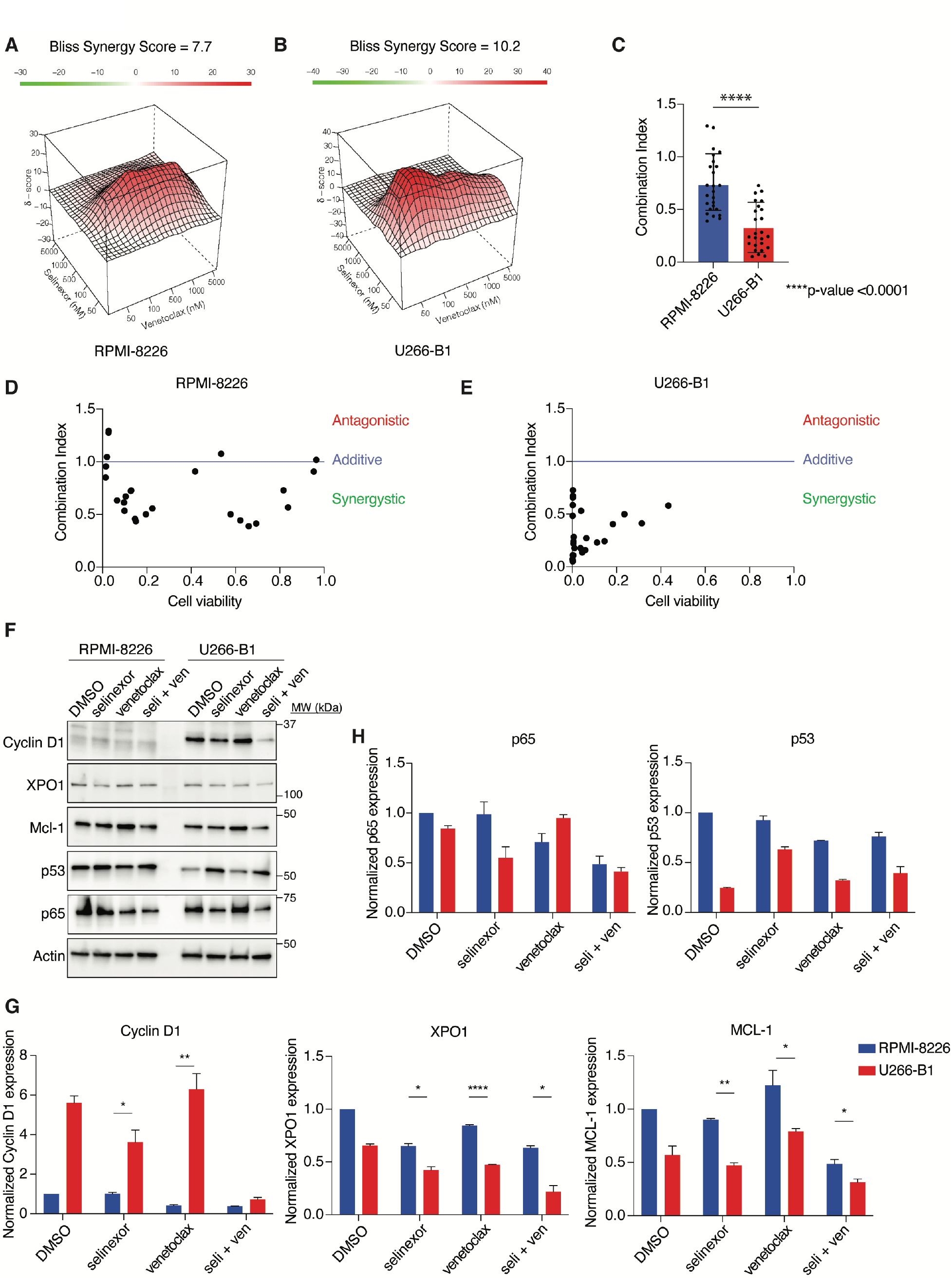
Combination of selinexor and venetoclax shows significant synergy and leads to a decrease in Cyclin D1, XPO1, and MCL-1 protein levels in U266-B1. RPMI-8226 and U266-B1 were treated with increasing doses of selinexor and venetoclax for 72 hours and cell viability was measured using CellTiter Glo. Contour plots were calculated using the Bliss Independence model and were generated using the SynergyFinder web application. Red indicates synergism and green indicates antagonism (**Panel A and B**). The synergy of RPMI-8226 and U266-B1 was compared using Combination Index (CI) values. The difference was measured using Student’s t-test (**Panel C**). RPMI-8226 and U266-B1 synergy was also calculated using the CompuSyn software. Combination Index values >1 indicates antagonism, =1 indicates additivity, <1 indicates synergy (**Panel D and E**). RPMI-8226 and U266-B1 were treated with selinexor (200nM) and venetoclax (1µM) for 24 hours and subjected to a Western blot using various antibodies as indicated (**Panel F**). The normalized protein levels of Cyclin D1, XPO1, MCL-1, p65, and p53 was calculated by the intensity of the Western blot bands using Image J software (**Panel G and H**).

We performed a western blot to determine the key protein changes with treatment in RPMI-8226 and U266-B1 (**Figure 2F**). We found that the t(11;14) cell line, U266-B1, was far more sensitive to the combination treatment of selinexor and venetoclax than RPMI-8226. Given that Cyclin D1 is known to be overexpressed in t(11;14)^1,2^, we measured Cyclin D1 levels and found that in RPMI-8226 (non-t(11;14)) Cyclin D1 was not expressed with any of the treatments; however, in U266-B1 (t(11;14)) Cyclin D1 was overexpressed but was decreased with selinexor, and the reduction was enhanced with the combination treatment (**Figure 2G**). We then measured XPO1 protein levels because it is inhibited by selinexor and prior studies show decreased XPO1 expression after XPO1 inhibition.^20^ As expected, with selinexor we saw a reduction in XPO1 protein levels that was further reduced with the combination in U266-B1 but no difference in selinexor and combination was seen in RPMI-8226. We then assessed the effects of treatment on cargo proteins that are known to be regulated by XPO1 and found an increase in tumor-suppressor p53 levels and a decrease in p65 levels with the treatment of selinexor and combination in both cell lines (**Fig. 2H**). Treatment of the multiple myeloma cell with venetoclax showed an upregulation of MCL-1 but was mitigated with the combination of selinexor and venetoclax. This effect was greater in U266-B1 and could explain the increased synergy alongside the combinatorial effects on Cyclin D1. An additional western blot was performed with treatment in non-t(11;14) cell line, OPM2, and t(11;14) cell line, KMS12BM (**Figure 1F** in the **Supplementary Appendix**). Cyclin D1, MCL-1, and p65 levels all significantly decreased with the combination treatment (**Figure 1G-H** in **Supplementary Appendix**).

### Combination selinexor and venetoclax therapy leads to decreased tumor volume and increased survival in an *in vivo* xenograft mice model

The effect of the combination of selinexor and venetoclax therapy in patients with multiple myeloma with t(11;14) we described above may have been due to either single drug alone; however, the *in vitro* synergy led us to hypothesize that the combination effect would be greater than the individual agents *in vivo* as well. We therefore utilized a xenograft multiple myeloma mouse model of the t(11;14) myeloma cell line KMS12BM. After tumor engraftment, mice were randomized to receive vehicle, selinexor, venetoclax or the combination of selinexor and venetoclax. Overall, the treatments were well tolerated, and the mice received continuous treatment until the vehicle recipients were euthanized due to advanced tumor growth at seventeen days. At this timepoint, the combination treated group showed a significant decrease in tumor volume when compared to the other groups (p <0.0001). There was a significant increase in overall survival of the mice in the combination treatment group compare to the other groups (p =0.0004; **Figure 3**).

**Figure 3.**
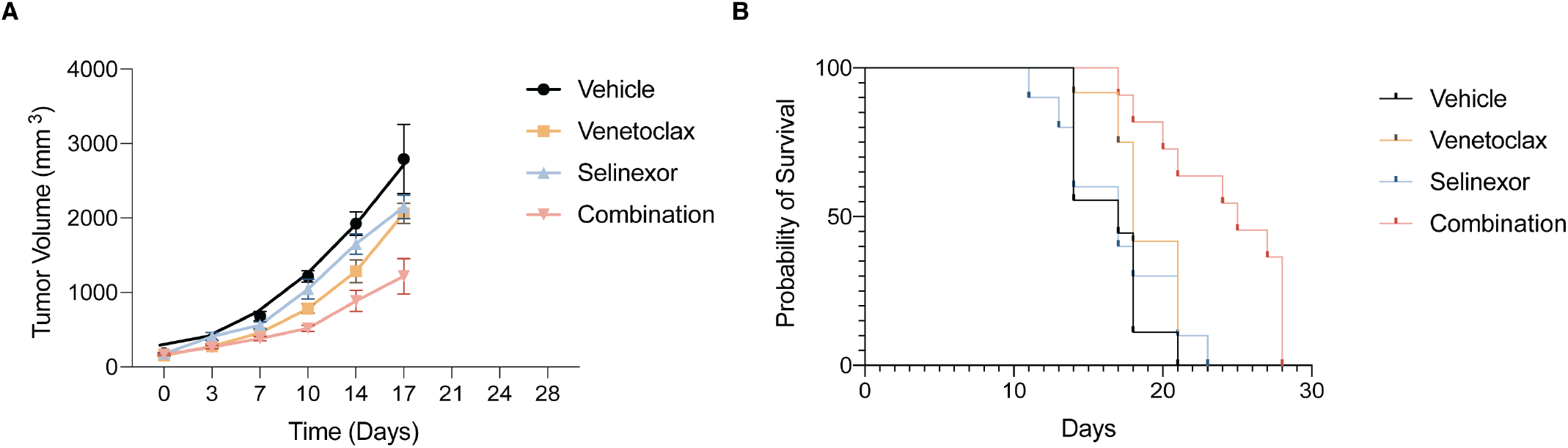
Combination of selinexor and venetoclax decreases tumor growth and increases survival in a mouse multiple myeloma xenograft model. Tumor volume measurements from mice randomized to receive treatment with vehicle, selinexor, venetoclax or the combination of selinexor and venetoclax (**Panel A**). The combination treated group showed a significant decrease in tumor volume when compared to the other groups (p=<0.0001). Mice were treated with selinexor (5mg/kg) three times per week, venetoclax (100mg/kg) daily, or the combination of the drugs on the same dose and schedule. Both drugs were given by oral gavage and vehicle treated mice were treated with both vehicles on the same schedule as the treated groups. The overall survival by Kaplan-Meier estimator is shown in **Panel B**. There was a significant increase in the survival of the combination treated group when compared to the other groups (p= 0.0004). Log rank test was used to compare survival.

## Discussion

In the past decade, the treatment landscape for MM has dramatically changed with proteasome inhibitors, immunomodulatory agents, and antibody therapies becoming the mainstays of therapy. Despite these innovations, cures for multiple myeloma remain elusive. However, successful management of the disease requires multiple lines of therapy and new drugs with novel mechanisms of action. Selinexor inhibits nuclear export with activity in penta-refractory MM. Venetoclax is approved in other hematologic malignancies and shows promise in a subset of MM patients with t(11;14). The combination of selinexor and venetoclax has shown preclinical synergy in other cancer types^21,22^ and is in Phase 1b clinical trials for relapsed, refractory Non-Hodgkin’s lymphoma or acute myeloid leukemia (NCT03955783; NCT04607772). To our knowledge, this is the first report of patients with MM treated with the combination of selinexor and venetoclax.

Four patients with t(11;14) MM with *CCND1*-*IGH* fusion confirmed by FISH who were relapsed or refractory to prior therapies were able to achieve responses with the combination of selinexor and venetoclax, with two patients achieving a VGPR. Importantly, both patients progressed through prior venetoclax-based regimens, yet they still responded to a selinexor and venetoclax combination. Neither patient received single agent selinexor, so we do not know whether the combination was required for the response. However, the doses of selinexor used were lower than that on the FDA label for use in refractory MM. Alongside the *in vitro* and *in vivo* synergy findings shown here, this suggests that there is an effect of the combination allowing for lower dosing of selinexor. Since adverse effects of selinexor have been shown to be dose-dependent, combining selinexor with venetoclax potentially allows for a more tolerable and therapeutic option for patients, such as the ones described, that are older, are heavily pre-treated, or not candidates for further intensive therapy. This may also serve as an option for patients who may not have access to clinical trials for relapsed and refractory disease or for certain reasons request a strictly oral regimen. It could also act as a bridge to therapies that require manufacturing time such as CAR-T therapy.

The synergistic mechanism of the combination and selective preference in t(11;14) multiple myeloma is an area of ongoing investigation. However, our preliminary studies suggest a role for Cyclin D1 itself as a target of selinexor and synergistic effect of the combination with venetoclax. Cyclin D1 is a cargo protein of XPO1, the direct target of selinexor, but the activity of venetoclax against Cyclin D1 expression is not well defined. Previous studies have correlated venetoclax sensitivity to the expression of BCL2, especially in cell lines or patients who possess t(11;14).^5,19,23^ Positive correlation between Cyclin D1 and BCL2 and the role in oncogenesis has been described in other tumor types.^24,25^ Our results did not show a difference in BCL2 levels after the combination therapy but other studies suggest the balance of BCL2 and other anti-apoptotic proteins, such as BCL-XL and MCL-1 against the pro-apoptotic proteins is more important than BCL-2 expression levels alone.^8,24,26–29^ Accordingly, we saw increases in MCL-1 levels with venetoclax monotherapy that was abrogated by the addition of selinexor. This effect was seen in both t(11;14) and non-t(11;14) myeloma cell lines but was more significantly decreased in the t(11;14). While MCL-1 has not been reported to be a direct cargo of XPO1, others have reported decreased MCL-1 levels by selinexor therapy potentially by disrupting protein and downregulated expression of mRNA.^30,31^ Expression of MCL-1 is also regulated by NF*κ*B signaling, and we observed decreases in NF*κ*B p65 after combination therapy.

In conclusion, though limited to four patients, we observed responses in four out of four patients the administration of selinexor and venetoclax combination was safe in t(11;14) translocated relapsed and refractory MM patients. Preclinical studies in a xenograft mouse model of t(11;14) multiple myeloma also showed combination efficacy and tolerability. Based on these results we are planning a prospective clinical trial to test the effectiveness of venetoclax and selinexor combination in t(11;14) multiple myeloma patients.

## Methods

### Patients

Patients received treatment under regular clinical care but retrospectively consented to be included in this study. The Sylvester Comprehensive Cancer Center (SCCC) institutional review board waived review of the study. Responses were assessed by IMWG criteria.^32^

### Animals

Three-month-old female NSG-SGM3 mice were used for xenograft studies approved by the University of Miami Institutional Animal Care and Use Committee (IACUC) guidelines (Protocol #20-079).

### Cell lines

RPMI-8226 and U266-B1 cell lines were purchased from ATCC (Manassas, VA). OPM2 and KMS12BM cell lines were a kind gift from Dr. Leif Bergsagel from Mayo Clinic. Cell lines were tested for mycoplasma with the MycoAlert mycoplasma detection kit (Catalog # LT07-218, Lonza, Morristown, NJ). RPMI-8226 and OPM2 were cultured in RPMI-1640 with 10% FBS. U266-B1 and KMS12BM were cultured in RPMI-1640 with 15% FBS and 20% FBS respectively. Cell lines were maintained at 37°C in a CO2 incubator and passaged every 2-3 days. Selinexor (Catalog # S7252, KPT-330) and venetoclax (Catalog # S8048, ABT-199) were obtained from Selleck Chemicals (Houston, TX).

### Cell Viability Assays

Cell lines were seeded at 1 × 10^4^ cells/well in a 6×6 matrix of white, clear-bottom 96 well plates. Each cell line was exposed to increasing concentrations of each drug and a vehicle-only control (DMSO). After 72 hours of incubation at 37°C in a 5% CO2 incubator, the effects of cell viability were measured using CellTiter-Glo viability assay according to the manufacturer’s instructions (Catalog # G7573, Promega, Madison, WI). Synergy was analyzed via the Bliss independence model using Synergy Finder software (synergy.fimm.fi) as well as via the Chou-Talalay method by using CompuSyn software (combosyn.com). Synergy Finder model synergy score values above 10 indicates synergistic effects and values below -10 indicates antagonistic drug effects. CompuSyn combination index (CI) values CI<1 are synergistic, CI=1 is additive, and CI>1 are antagonistic.

### Immunoblotting

Cells were plated at 1 × 10^6^ cells/well in a 6-well plate. Whole cell lysates were prepared with IP lysis buffer supplemented with protease and phosphatase inhibitors (Thermo Scientific, Waltham, MA). Protein concentration was determined using BCA Protein Assay kit (Thermo Scientific, Waltham, MA). Ten micrograms of total protein was separated by electrophoresis on a 4-12% bis-tris protein gel and transferred onto PVDF membranes and probed with antibodies against Cyclin D1 (BD Biosciences, San Jose, CA); XPO1 and p53 (Santa Cruz Biotechnology, Dallas, TX); and MCL-1 and p65 (Cell Signaling Technologies, Danvers, MA), all used at 1:1000 dilution in 1% BSA. β-actin (Sigma Aldrich) was used at 1:10,000. Membranes were visualized by Clarity Western ECL substrate (BioRad, Hercules, CA) following the manufacturer’s protocol. The following treatments were used: DMSO-only control, selinexor (200nM), venetoclax (1µM), and combination. β-actin was used as a loading control. Treatment of U266-B1 and RPMI-8226 was 24 hours and treatment of KMS12BM and OPM2 was 16 hours. Western blot quantification was performed using ImageJ and normalized against β-actin.

### Xenograft studies and in vivo treatments

The NSG-SGM3 mice were weighed followed by anesthetized by exposure to 1-5% vaporized isoflurane in 100% oxygen. One million KMS12BM cells were injected into the mice subcutaneously. The following drugs were administered with a minimum of nine mice per experimental group: vehicle on the same schedule as the treated groups, selinexor (5mg/kg; oral gavage three times a week), venetoclax (100mg/kg; oral gavage daily), and the combination on the same schedule. Mice were monitored daily, and tumor burden was measured using high-frequency ultrasound (Vevo3100, Visualsonics). Mice were sacrificed when tumor size reached >2mm^3^ or if weight loss was >20% for two consecutive days.

## Data Availability

All data produced in the present work are contained in the manuscript

## Data Availability

All data will be made available by contacting the corresponding authors.

## Acknowledgements

We would like to thank the patients and their families.

## Author Contributions

*Conceptualization:* N.N., J.H., J.T.

*Investigation:* S.C., T.T., R.D., E.R., S.M., G.P., A.P., J.A., M.A., J.J., T.B, F.M, D.K, D.B., J.C.

*Writing-Original draft preparation:* N.N., S.C., J.T.

*Writing-Reviewing and Editing:* N.N., S.C., F.M., O.L., J.H., J.T.

## Competing Interests

**Bradley:** AbbVie: Membership on an entity’s Board of Directors or advisory committees, Speakers Bureau; Novartis: Consultancy, Membership on an entity’s Board of Directors or advisory committees. **Maura:** OncLive: Honoraria; Medscape: Consultancy, Honoraria. **Kazandjian:** Arcellx: Honoraria, Membership on an entity’s Board of Directors or advisory committees; BMS: Honoraria, Membership on an entity’s Board of Directors or advisory committees. **Landgren:** Adaptive: Honoraria; Binding Site: Honoraria; BMS: Honoraria; Cellectis: Honoraria; Amgen: Honoraria; Janssen: Honoraria; Celgene: Research Funding; Janssen: Other: IDMC; Janssen: Research Funding; Takeda: Other: IDMC; Amgen: Research Funding; GSK: Honoraria. **Taylor:** Karyopharm: Honoraria.

## Notes

### Funding Statement

This study was funded by grants from the U.S. Department of Health & Human Services | NIH | National Cancer Institute (NCI) - K08CA230319 [Taylor] and the Doris Duke Charitable Foundation (DDCF) - CSDA [Taylor]

### Author Declarations

The institutional review board of Sylvester Comprehensive Cancer Center waived ethical approval for this work.

