## Supplementary Appendix for "Combination Venetoclax and Selinexor Effective in Relapsed Refractory Multiple Myeloma with Translocation t(11;14)"

### Table of Contents:

**Supplementary Figure and Text (page 2-3)**
**Supplementary Table (page 4)**

Nina Nguyen, MD[1#], Sana Chaudhry, BSc[1#], Tulasigeri M. Totiger, PhD[1#], Robert Diaz, MS[1], Evan Roberts, BSc[1], Skye Montoya, BSc[1], Gabriel Pardo BSc[1], Alejandro Pardo BSc[1], Jumana Afaghani, BSc[1], Maurizio Affer, MSc[1], Jacob Jahn, BSc[1], Terrence Bradley, MD[3], Francesco Maura, MD[2], Dickran Kazandjian, MD[2], Daniel Bilbao, PhD[1], Jennifer Chapman, MD[4], Ola Landgren, MD,PhD[2], James Hoffman, MD[2*], and Justin Taylor, MD[3*]

### Supplementary Figure 1

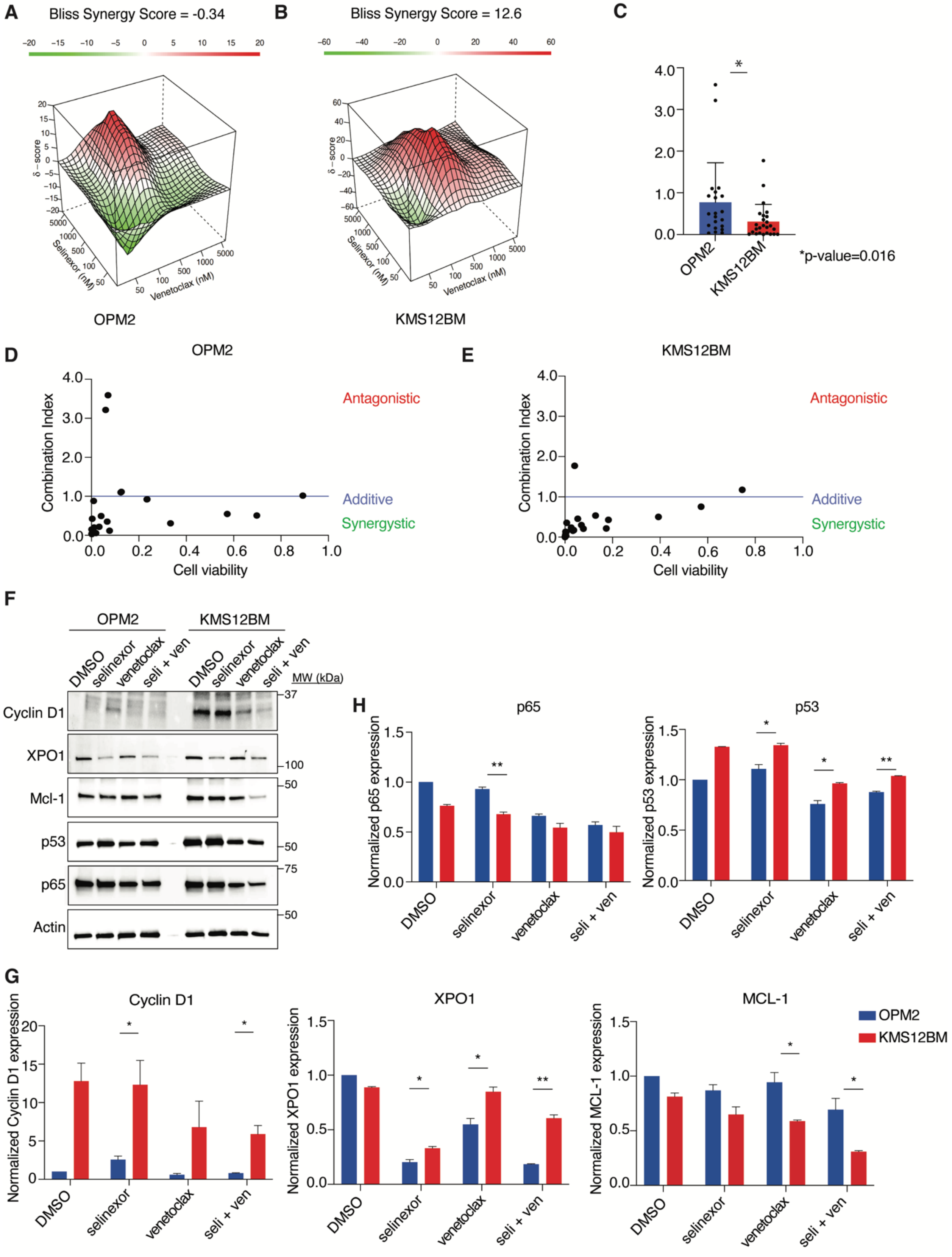

**Supplementary Figure 1. The combination of selinexor and venetoclax show higher levels of synergy and decreased levels of Cyclin D1, XPO1, and MCL-1 protein levels in KMS12BM.**

OPM2 and KMS12BM were treated with increasing doses of selinexor and venetoclax for 72 hours and cell viability was measured using CellTiter-Glo. Contour plots were calculated using the Bliss Independence model and were generated using the SynergyFinder web application. Red indicates synergism and green indicates antagonism (**Panel A and B**). The synergy of OPM2 and KMS12BM was compared using Combination Index (CI) values. The difference was measured using Student's t-test (**Panel C**). The synergy was also calculated using the CompuSyn software. Combination Index values >1 indicates antagonism, =1 indicates additivity, <1 indicates synergy (**Panel D and E**). OPM2 and KMS12BM were treated with selinexor (200nM) and venetoclax(1μM) for 16 hours and subjected to a Western blot using various antibodies as indicated (**Panel F**). The normalized protein levels of p65, p53, Cyclin D1, XPO1, and MCL-1 was calculated by the intensity of the Western blot bands using Image J software (**Panel G and H**).

| Cell Lines | MM IgH translocation |
|---|---|
| U266-B1 | t(11;14)(CCND1/IGH) |
| KMS-12-BM | t(11;14)(CCND1/IGH) |
| MOLP-8 | t(11;14)(CCND1/IGH) |
| SK-MM-2 | t(11;14)(CCND1/IGH) |
| NCI-H929 | t(4;14)(MMSET/IGH) |
| LP-1 | t(4;14)(MMSET/IGH) |
| OPM-2 | t(4;14)(MMSET/IGH) t(?;20)(?;MAFB) |
| EJM | t(14;20)(IGH/MAF) |
| L-363 | t(6;20)(?;MAFB) |
| KARPAS-620 | t(8;11)(CCND1) |
| JJN-3 | t(14;16)(IGH/MAF) |
| MM1S | t(14;16)(IGH/MAF) |
| KMS-11 | t(4;14)(MMSET/IGH) t(14;16)(IGH/MAF) |
| RPMI-8226 | t(16:22)(MAF;IGL) |
| AMO-1 | - |
| MC-CAR | - |
| IM-9 | - |
| ARH-77 | - |

**Supplemental Table 1. List of multiple myeloma (MM) cell lines according to their immunoglobulin heavy chain (IgH) translocation.**

Abbreviations: CCND1, Cyclin D1; IGH, immunoglobulin heavy chain; MMSET, multiple myeloma SET domain; IGL, immunoglobulin light chain
